# Deep learning Enables Accurate Diagnosis of Novel Coronavirus (COVID-19) with CT images

**DOI:** 10.1101/2020.02.23.20026930

**Authors:** Song Ying, Shuangjia Zheng, Liang Li, Xiang Zhang, Xiaodong Zhang, Ziwang Huang, Jianwen Chen, Huiying Zhao, Ruixuan Wang, Yutian Chong, Jun Shen, Yunfei Zha, Yuedong Yang

**Affiliations:** National Supercomputer Center in Guangzhou, Sun Yat-sen University, Guangzhou 510006, China; Department of Radiology, Renmin Hospital of Wuhan University, Wuhan 430060, China; Department of Radiology, Sun Yat-Sen Memorial Hospital, Sun Yat-Sen University, Guangzhou 510530, China; Department of Radiology, The Third Affiliated Hospital of Sun Yat-Sen University, Guangzhou 510220, China; School of Data and Computer Science, Sun Yat-sen University, Guangzhou 510006, China; School of Systems Sciences and Engineering, Sun Yat-sen University, Guangzhou 510006, China

## Abstract

**Background:** A novel coronavirus (COVID-19) has emerged recently as an acute respiratory syndrome. The outbreak was originally reported in Wuhan, China, but has subsequently been spread world-widely. As the COVID-19 continues to spread rapidly across the world, computed tomography (CT) has become essentially important for fast diagnoses. Thus, it is urgent to develop an accurate computer-aided method to assist clinicians to identify COVID-19-infected patients by CT images.

**Materials and Methods:** We collected chest CT scans of 88 patients diagnosed with the COVID-19 from hospitals of two provinces in China, 101 patients infected with bacteria pneumonia, and 86 healthy persons for comparison and modeling. Based on the collected dataset, a deep learning-based CT diagnosis system (DeepPneumonia) was developed to identify patients with COVID-19.

**Results:** The experimental results showed that our model can accurately identify the COVID-19 patients from others with an excellent AUC of 0.99 and recall (sensitivity) of 0.93. In addition, our model was capable of discriminating the COVID-19 infected patients and bacteria pneumonia-infected patients with an AUC of 0.95, recall (sensitivity) of 0.96. Moreover, our model could localize the main lesion features, especially the ground-glass opacity (GGO) that is of great help to assist doctors in diagnosis. The diagnosis for a patient could be finished in 30 seconds, and the implementation on Tianhe-2 supercompueter enables a parallel executions of thousands of tasks simultaneously. An online server is available for online diagnoses with CT images by http://biomed.nscc-gz.cn/server/Ncov2019.

**Conclusions:** The established models can achieve a rapid and accurate identification of COVID-19 in human samples, thereby allowing identification of patients.

## Introduction

In late December, 2019, a cluster of pneumonia cases with unknown etiology were reported in Wuhan city, Hubei Province, China.^1^ Deep sequencing analyses from lower respiratory tract samples revealed a novel coronavirus that was currently named 2019 novel coronavirus (COVID-19) by the World Health Organization (WHO),^2^ which resembled severe acute respiratory syndrome coronavirus (SARS-CoV).^3^ Outbreaks in healthcare workers and families indicated human-to-human transmission.^1,4^ The COVID-19 is a beta-CoV of group 2B with at least 70% similarity in genetic sequence to SARS-CoV ^5^ and is the seventh member of the family of enveloped RNA coronavirus that infect humans. ^6^

Since January 17, the confirmed cases dramatically increased, and COVID-19 has been designated as a public health emergency of international concerns by the WHO. As of February 23, 2020, China has documented 77048 confirmed cases while the death toll rose to 2445. Moreover, more than 2000 confirmed cases were identified in other countries worldwide. The COVID-19 has posed significant threats to international health. ^7^

The spectrum of this disease in humans is yet to be fully determined. Signs of infection are nonspecific, including respiratory symptoms, fever, cough, dyspnea, and viral pneumonia.^1^ With the daily increase in the number of newly diagnosed and suspected cases, the diagnosis has become a growing problem in major hospitals due to the insufficient supply of nucleic acid detection boxes and limited detection rates in the epidemic area.^8^ Computed tomography (CT) and radiography have thus emerged as integral players in the COVID-19’s preliminary identification and diagnosis.^9-11^ However, the overwhelming patients and relatively insufficient radiologists led to high false positive rate.^11^ Advanced computer-aided lung CT diagnosis systems are urgently needed for accurately confirming suspected cases, screening patients, and conducting virus surveillance.

In this study, we have developed a deep learning-based lung CT diagnosis system to detect the patients with COVID-19, which can automatically extract radiographic features of the novel pneumonia, especially the ground-glass opacity (GGO) from radiographs.

## Materials and Methods

Our institutional review board waived written informed consent for this study that evaluated de-identified data and involved no potential risk to patients.

### Data acquisition

This study is based on the reliable resources that were provided by the Renmin Hospital of Wuhan University, and two affiliated hospitals (the Third Affiliated Hospital and Sun Yat-Sen Memorial Hospital) of the Sun Yat-sen University in Guangzhou.

We obtained totally CT images of 88 COVID-19 infected patients, which comprised of 76 and 12 patients from the Renmin Hospital of Wuhan University and the Third Affiliated Hospital, respectively. Wuhan’s chest CT examinations were performed with a 64-section scanner (Optima 680, GE Medical Systems, Milwaukee, WI, USA) without the use of contrast material. The CT protocol was as follows: 120 kV; automatic tube current; detector, 35 mm; rotation time, 0.35 second; section thickness, 5 mm; collimation, 0.75 mm; pitch, 1-1.2; matrix, 512×512; and inspiration breath hold. The images were photographed at lung (window width, 1000–1500 HU; window level, –700 HU) and mediastinal (window width, 350 HU; window level, 35– 40 HU) settings. The reconstructions were made at 0.625 mm slice thickness on lung settings. These data consist of only transverse plane images of lung. For patients in Guangzhou, we obtained his/her lung images in anterior view, lateral view and transverse view. The chest CT examinations were performed with a 64-slice spiral scanner (Somatom Sensation 64; Siemens, Germany).

All patients’ nasopharyngeal swabs were subjected to nucleic acid kit lysis extraction and calculation to the laboratory of Renmin Hospital of Wuhan University performed fluorescence RT-PCR to detect the viral nucleic acid gene sequence, and compared it with the novel coronavirus nucleocapsid protein gene (nCoV-NP) and the novel coronavirus open reading coding frame lab (nCoV ORFlab) sequence. The results were positive, and the HRCT image quality of the chest was acceptable, with no significant artifacts or missing images.

For comparison, we also retrieved CT underwent chest images of 86 healthy people and 100 patients with bacterial pneumonia from the Renmin Hospital of Wuhan University and Sun Yat-Sen Memorial Hospital.

### Model Architecture and Model Training

We developed a deep learning-based CT diagnosis system (referred to as DeepPneumonia) to assist doctors to detect the COVID-19 causing pneumonia and to localize the main lesions. As shown in Figure 1, our fully automated lung CT diagnosis system was developed by three main steps. First, we extracted the main regions of lungs and filled the blank of lung segmentation with the lung itself to avoid noises caused by different lung contours. Then, we designed a Details Relation Extraction neural network (DRE-Net) to extract the top-K details in the CT images and obtain the image-level predictions. Finally, the image-level predictions were aggregated to achieve patient-level diagnoses.

**Figure 1:**
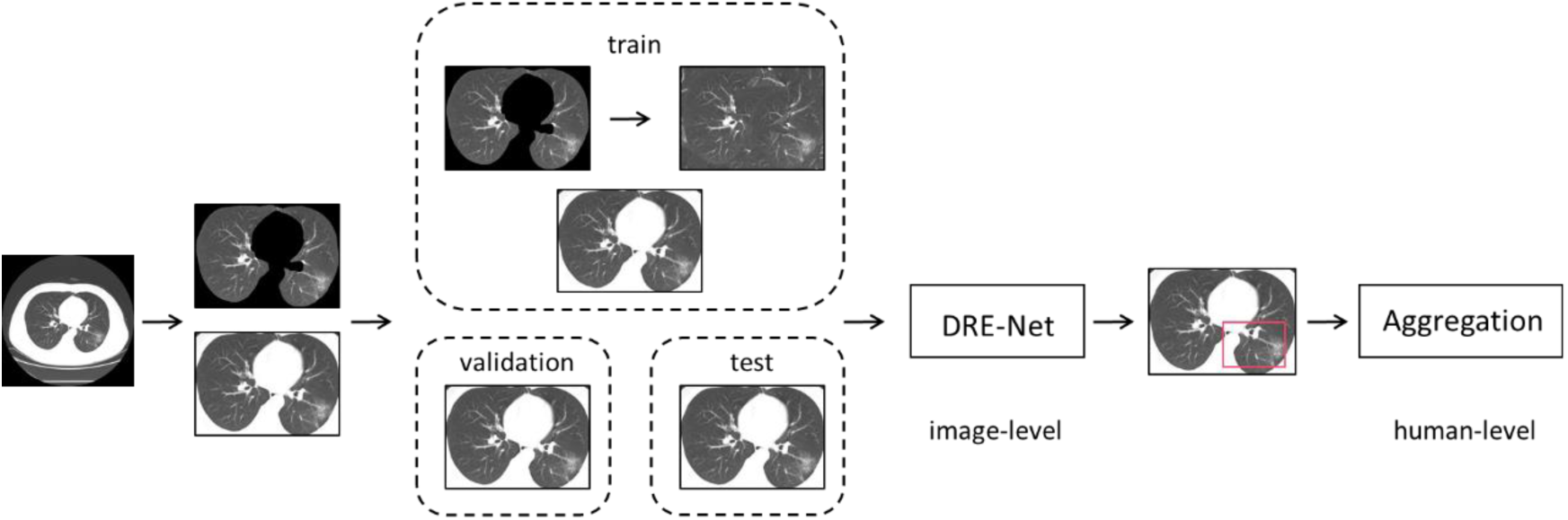
Illustration of DeepPneumonia framework.

### Data preprocessing

Each set of 3D CT images was equally divided into 15 slices. The slices with incomplete lung were removed. The lung region in each slice were automatically extracted by the open source package OpenCV.^12^ As the lung contours are of large differences between humans, the images were filled with an background composed of 10 translational and rotational lungs. Finally, we kept 88 COVID-19 patients with 777 CT images, 100 bacterial pneumonia patients with 505 slices, and 86 healthy people with 708 slices in this study.

### DRE-Net

Our training models were concretely constructed on the pretrained ResNet-50^13^, on which the Feature Pyramid Network (FPN) ^14^ was added to extract the top-K details from each image. An attention module is coupled to learn the importance of each detail.^15^ By using the FPN and attention modules, our model can not only detect the most important part of images, but also interpret the outputs by the neural network.

### Aggregation

The image-level scored results of slices were aggregated for each patient. Here, the mean pooling was used to integrate the image-level results into the morbidity of each person as the human-level result.

### Implementation and evaluation

For practical purposes, we designed the models with two tasks, discriminating COVID-19-infected patients from the bacterial-infected patients, and separating COVID-19 patients from healthy controls, respectively. For each task, we employed the patient-level split strategy following the LUNA16 competition^16^ by using random splits of 60%/10%/30% for training, validation, and test sets, respectively. The training set were used to train models, and the validation set was used to optimize the hyperparameters for the best performance. The final optimal models were independently assessed on the test set. We computed accuracy, precision, recall as

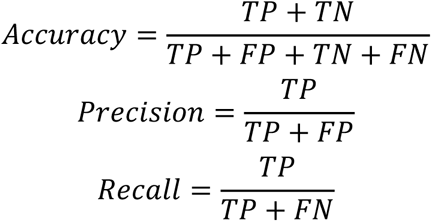

where TP, TN, FP, and FN are the numbers of true positives, true negatives, false positives, and false negatives, respectively. We also computed Area under the receiver-operating characteristics Curve (AUC) by the scikit-learn 0.19 (scikit-learn.org).

## Results and discussion

### Deep Learning Model for Pneumonia Classification

An important tool for assisted diagnoses is to accurately discriminate the bacterial pneumonia and viral pneumonia (COVID-19) in order to decrease the unnecessary waste of medical resources. To this end, we built a pneumonia classification model based on 88 COVID-19 and 100 bacterial pneumonia patients, from which we collected 777 and 505 images. By tuning hyperparameters according to the validation set, we achieved AUC of 0.91 in the image level, and AUC of 0.95 in the human level. The performances are essentially the same on the test set with AUC of 0.92 in the image level, and AUC of 0.95 and recall of 0.96 in the human level. The higher AUC in the test set is partly due to the relatively small datasets. To indicate the effectiveness of our proposed DRENet architecture, we also employed deep residual network (Resnet),^13^ DenseNet^17^, and VGG16^18^ for comparisons. As shown in Table 1, by selecting the top-K important details and the relationships among them, our DRE-Net improves about 7.0% recall and 3.0% F1-score over VGG16 with the same fold assignments, respectively. Figure 2 shows the receiver-operating characteristic curves for comparisons with other baseline models and the confusion matrix of DRE-Net on the test set.

**Table 1.** Performance comparisons on pneumonia classification dataset.

| Model | AUC | Recall | Precision | F1-score | Accuracy |
| --- | --- | --- | --- | --- | --- |
| VGG16 | 0.91 | 0.89 | 0.80 | 0.84 | 0.84 |
| DenseNet | 0.87 | 0.93 | 0.76 | 0.83 | 0.82 |
| ResNet | 0.90 | 0.93 | **0.81** | 0.86 | 0.86 |
| **DRE-Net** | **0.95** | **0.96** | 0.79 | **0.87** | **0.86** |

**Figure 2:**
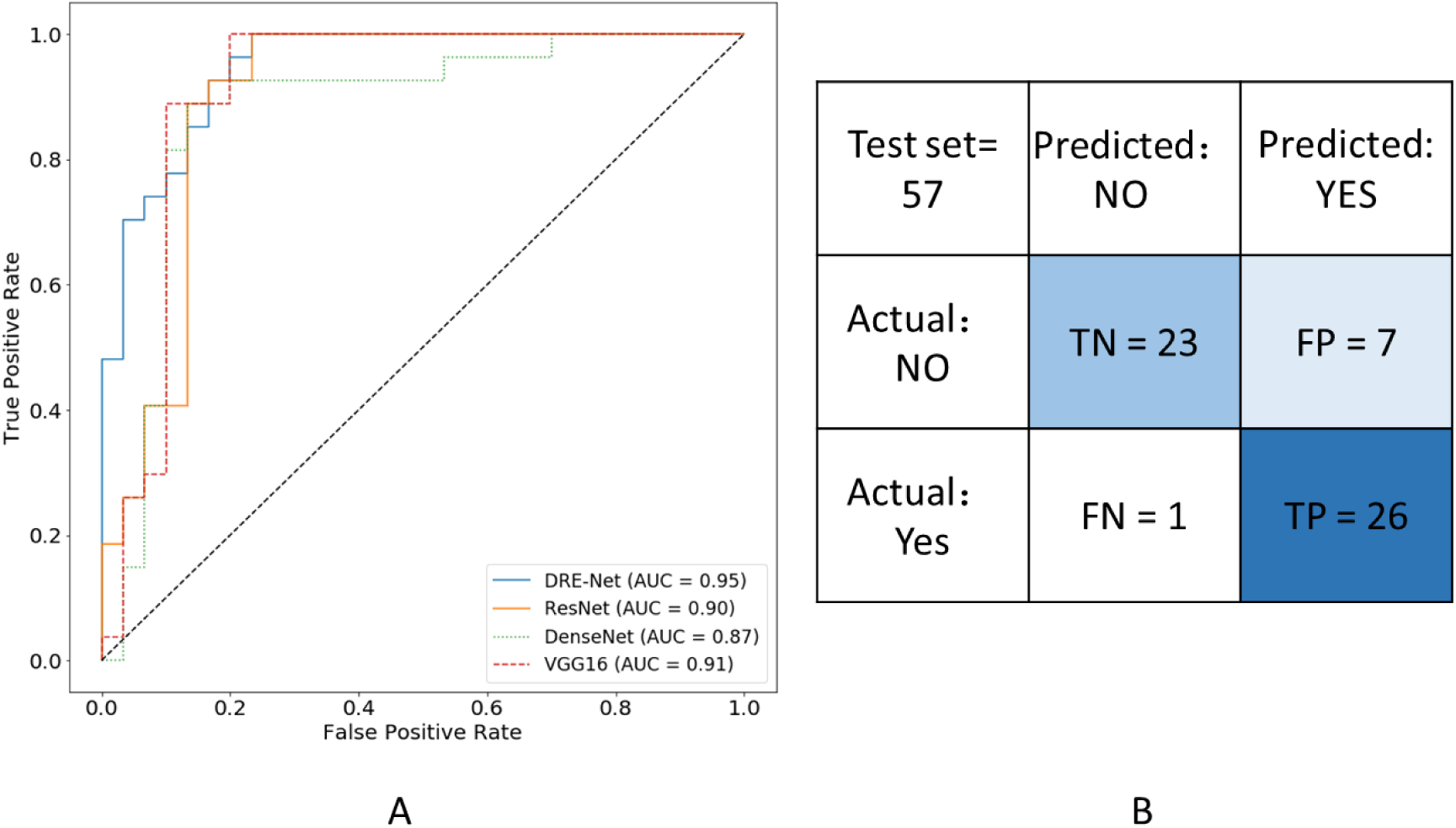
Performance of the DRE-Net on the pneumonia classification. A) Receiver operating characteristic curves for the classification between COVID-19 and normal bacteria pneumonia. B) Confusion matrix of the DRE-Net on the test set.

### Deep Learning Model for Pneumonia Diagnosis

Additionally, we also performed experiments on 88 COVID-19 patients and 86 healthy persons, from which 777 and 708 images were collected. By tuning hyperparameters on the 10% validation set, we achieved AUC of 0.97 in the image level, and 0.99 in the patient level. The performances are essentially the same on the test set with 0.98 in the image level, and AUC of 0.99 and recall of 0.93 in the patient level.

As shown in Table 2, DRE-Net achieved better performance than those of ResNet, DenseNet and VGG16 because of the strong power of details extraction. The high performance of these deep learning models also demonstrated that the CT images of pneumonia patients and healthy people were well differentiated.

**Table 2.** Performance comparisons on pneumonia diagnosis dataset.

| Model | AUC | Recall | Precision | F1-score | Accuracy |
| --- | --- | --- | --- | --- | --- |
| VGG16 | 0.98 | 0.89 | 0.92 | 0.91 | 0.90 |
| DenseNet | 0.97 | 0.92 | 0.85 | 0.92 | 0.92 |
| ResNet | 0.99 | 0.89 | 0.96 | 0.92 | 0.92 |
| **DRE-Net** | **0.99** | **0.93** | **0.96** | **0.94** | **0.94** |

The receiver-operating characteristic curves for comparisons with other baseline models and the confusion matrix are shown in Figure 3.

**Figure 3:**
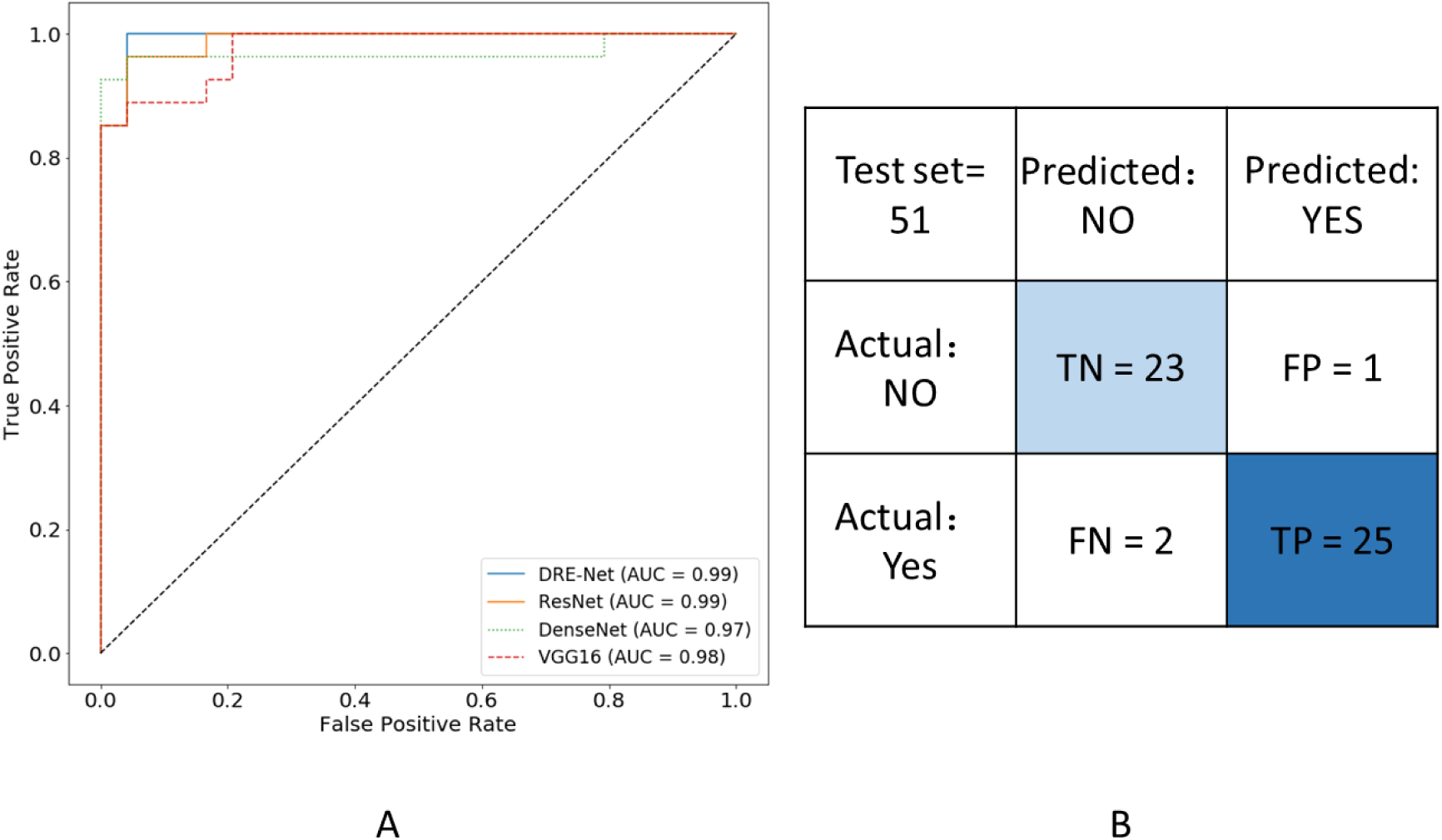
Performance of the DRE-Net on the pneumonia diagnosis. A) Receiver operating characteristic curves for the diagnosis of COVID-19. B) Confusion matrix of the DRE-Net on the test set.

### Model Interpretation and Visualization

Another advantage of our model is its interpretability. In diagnosis of COVID-19, ground-glass opacity (GGO) in CT image is one of the most important factors to recognize the patients.^9-11^ Therefore, we expected our model can localize GGO in CT images, especially in the early stage patients and suspected patients. We visualized the automatically extracted details of two successfully predicted pneumonia patients from the test set. For each patient, the top 3 predicted slices and the extracted details were indicated with predicted scores above 0.8 (range from 0 to 1). As shown in Figure 4, the assessments of DRNet mainly focused on the region of the GGO abnormality. These findings indicate that DRNet learned to assess the correct features instead of learning image correlations. Moreover, our model provides reasonable clues on the factors for its judgements, which is of great help to assist doctors in diagnosis.

**Figure 4:**
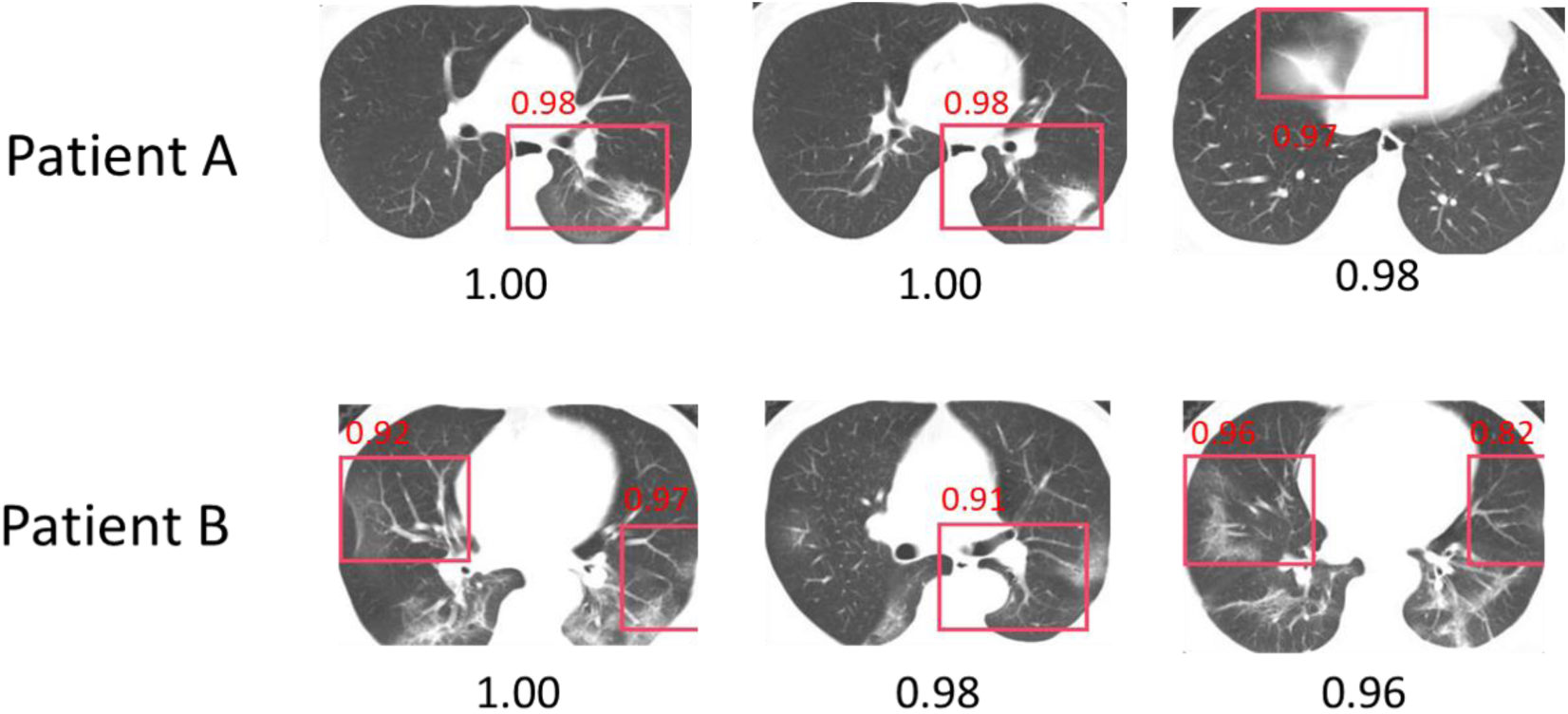
Visualization of the DeepPneumonia on the pneumonia diagnosis. For each patient, we showed the top-3 predicted slices and the extracted details (bounding boxes with red color) with normalized predicted scores above 0.8 (range from 0 to 1). The model can accurately identify the regions of the GGO abnormality.

## Conclusion

In conclusion, our study demonstrated the feasibility of a deep learning approach to assist doctors to detect the patients with COVID-19 and automatically to identify the lesions from CT images. By achieving a high performance on both the pneumonia detection and classification tasks, the proposed system may enable a rapid identification of patients.

## Data Availability

To avert any potential breach of confidentiality, no link between the patients and the researchers were made available.

http://biomed.nscc-gz.cn/server/Ncov2019

## Funding

The work was supported in part by the National Key R&D Program of China (2018YFC0910500), National Natural Science Foundation of China (U1611261, 61772566, and 81801132), Guangdong Frontier & Key Tech Innovation Program (2018B010109006 and 2019B020228001), Natural Science Foundation of Guangdong, China (2019A1515012207), and Introducing Innovative and Entrepreneurial Teams (2016ZT06D211).

## Notes

### Competing Interest Statement

The authors have declared no competing interest.

### Clinical Trial

No clinical trials involve

